# Electrical Brain Stimulation and Continuous Behavioral State Tracking in Ambulatory Humans

**DOI:** 10.1101/2021.08.10.21261645

**Authors:** Filip Mivalt, Vaclav Kremen, Vladimir Sladky, Irena Balzekas, Petr Nejedly, Nick Gregg, Brian Lundstrom, Kamila Lepkova, Tereza Pridalova, Benjamin H. Brinkmann, Pavel Jurak, Jamie J. Van Gompel, Kai Miller, Timothy Denison, Erik St Louis, Gregory A. Worrell

## Abstract

**Objective:** Electrical deep brain stimulation (DBS) is an established treatment for patients with drug-resistant epilepsy. Sleep disorders are common in people with epilepsy, and DBS may actually further disturb normal sleep patterns and sleep quality. Novel devices capable of DBS and continuous intracranial EEG (iEEG) telemetry enable detailed assessments of therapy efficacy and tracking sleep related comorbidities. Here, we investigate the feasibility of automated sleep classification using continuous iEEG data recorded from Papez’s circuit in four patients with drug resistant mesial temporal lobe epilepsy using an investigational implantable sensing and stimulation device with electrodes implanted in bilateral hippocampus (HPC) and anterior nucleus of thalamus (ANT).

**Approach:** The iEEG recorded from HPC is used to classify sleep during concurent DBS targeting ANT. Simultaneous polysomnography and HPC sensing were used to train, validate and test an automated classifier for a range of ANT DBS frequencies: no stimulation, 2 Hz, 7 Hz, and high frequency (>100 Hz).

**Main results:** We show that it is possible to build a patient specific automated sleep staging classifier using power in band features extracted from one HPC sensing channel. The patient specific classifiers performed well under all thalamic DBS frequencies with an average F1-score 0.894, and provided viable classification into awake and major sleep categories, rapid eye movement (REM) and non-REM. We retrospectively analyzed classification performance with gold-standard polysomnography annotations, and then prospectively deployed the classifier on chronic continuous iEEG data spanning multiple months to characterize sleep patterns in ambulatory patients living in their home environment.

**Significance:** The ability to continuously track behavioral state and fully characterize sleep should prove useful for optimizing DBS for epilepsy and associated sleep, cognitive and mood comorbidities.

## 1. Introduction

Electrical deep brain stimulation (DBS) is an established therapy for drug-resistant focal epilepsy [1–4], but the effects of DBS on sleep are poorly understood. A small study of responsive neurostimulation (RNS) with only hippocampal or cortical stimulation did not find evidence for sleep disruption [5], but thalamic DBS targeting anterior nucleus of thalamus (ANT) has been reported to cause sleep disruption [6].

With the high prevalence of sleep disturbances in people with epilepsy [7–12], and the potential for seizures to follow sleep related circadian patterns [13–16], the precise impact of DBS on sleep is of significant clinical importance. Morevoer, sleep is known to play an important role in the cognitive [8,17] and mood [18,19] comorbidities of epilepsy. Unfortunately, objectively assessing sleep in patients undergoing DBS has been difficult due to the absence of longitudial data and the unreliability of patient sleep self reporting [20]. A quantiative assessment of sleep architecture using ambulatory intracranial EEG (iEEG) data in patients with epilepsy allows investigation of long-term behavioral state dynamics and the complex interplay of epilepsy, sleep and DBS that should ultimately increase the precision of adaptive DBS [21].

Although clinical gold standard polysomnography involves trained experts reviewing scalp EEG recordings, prior research has shown the feasibility of automated behavioral state classification utilizing invasively recorded iEEG signals. These studies demonstrated the feasibility of classification of wakefulness and non-Rapid-Eye-Movement (non-REM: N2 & N3) sleep stages [22–25]. However, the feasibility of iEEG based REM classification, and the impact of DBS induced iEEG artifacts on automated sleep scoring remains unclear.

Our understanding of the chronic effects of DBS on sleep has been limited by the recording capabilities of current DBS devices [26–28]. Recent advances in DBS devices have included rechargeable batteries that support the energy demands of continuous iEEG data telemetry. One such system, the investigational Medtronic Summit RC+S™, currently in use under an investigational device exemption enables continuous iEEG streaming during therapeutic DBS [28,29]. With continuous iEEG streaming and bidirectional connectivity the investigational Medtronic Summit RC+S™ system provides a unique opportunity for long-term, ambulatory monitoring and quantitative evaluation of sleep during DBS therapy [28,29].

We collected ambulatory iEEG recordings from four patients with drug resistant epilepsy implanted with the investigational Medtronic Summit RC+S™ to investigate novel stimulation paradigms and to track long term behavioral state dynamics. The patients received therapeutic ANT DBS during concurrent, bilateral hippocampal (HPC) iEEG recording. We evaluated the feasibility and accuracy of automated behavioral state classification under different ANT stimulation frequencies (2 Hz, 7 Hz, high frequency >100 Hz) [2,30–34] during three days of simultanous iEEG and polysomnography (PSG) with expert sleep annotations. A Naïve Bayes classifier [35,36] was used for classifying iEEG signals into Awake, Rapid-Eye-Movement (REM), and non-REM (non-REM: N2 & N3). Subsequently, we deployed the trained classifiers in four ambulatory patients over 6 months.

Previous studies investigating automated behavioral state classification with iEEG have not explored the impact of concurrent DBS on classifier performance, and have only utilized semi-gold standard annotations [22], or performed leave-out cross validation testing [24]. Furthermore, to our knowledge automated behavioral state classifiers have not been deployed in ambulatory subjects living in their natural environments. Here we obtained simultaneous polysomnography and iEEG during both low- and high-frequency ANT DBS using a novel investigational implantable neural sensing and stimulation device to create gold-standard behavioral state labels for training, validation and testing of automated behavioral state classifiers that were then deployed in ambulatory subjects during therapeutic ANT DBS.

## 2. Methods

### 2.1 Experimental Protocol

This human subjects research study was carried out under a Food and Drug Administration investigational device exemption (IDE-G180224) and Mayo Clinic institutional review board (IRB: 18-005483 “Human Safety and Feasibility Study of Neurophysiologically Based Brain State Tracking and Modulation in Focal Epilepsy”) approvals. The study is registered at https://clinicaltrials.gov/ct2/show/NCT03946618. The patients provided written consent in accordance with the IRB and FDA requirements. Four subjects with drug resistant mesial temporal lobe epilepsy (mTLE) (H1-4) were consented and implanted with the investigational Medtronic Summit RC+S™ device [29,37,38], with four 4-contact leads, targetting the HPC and ANT) bilaterally. See supplementary data for additional surgical and clinical information.

We collected simultaneous polysomnography and iEEG data in the hospital setting to obtain gold standard sleep classifications based on expert review of scalp EEG (EKS) for subsequent traning, validation and testing of the iEEG-based automated classifier. Subsequently, continuous prospective automated classifications were made in naturalistic settings in ambulatory patients, without concurrent scalp recordings.

The overnight hospital and long-term experiments were conducted within a broader clinical study in temporal lobe epilepsy investigating the effect of multiple stimulation frequencies on interictal epileptiform discharges and seizures. The stimulation parameters were selected based on current clinical practice and experimental studies investigating the therapuetic effect of ANT DBS [2,30–34]. The HPC sensing parameters, including sampling freqency, were determined as a compromise between data quality and battery life-cycle duration.

#### 2.1.1 Intracranial EEG Data Acquisition

Intracranial EEG data were continuously collected using wireless streaming to a tablet computer with 250 Hz or 500 Hz sampling frequency as previously described [37–39]. Data acquired at 500 Hz was downsampled to 250 Hz using an anti-aliasing finite response filter (101^th^ order) with cutoff frequency at 100 Hz. Impedance between the implanted device and electrode contacts was below 2 kΩ for all contacts during all experiments. The long term iEEG data from four patients spanned over 1,000 days, and here we analyzed at least 30 days for each subject, H1-4.

#### 2.1.2 Simultaneous Polysomnography (PSG) and Intracranial EEG (iEEG) Streaming During Electrical Brain Stimulation

Simultaneous HPC iEEG and PSG (scalp EEG, eye leads, chin leads) recording was conducted at different ANT DBS parameters (no DBS, 2 Hz, 7 Hz and high frequency (>100 Hz) DBS; 3 – 4 mA; 90 and 200 µs pulse width) over the course of three consecutive nights in the hospital epilepsy monitoring unit. This provided standard reference labels for training, validation and testing of the behavioral state classifier. The Natus Medical Inc. Xltek electrophysiology system was used to acquire all PSG data with a common reference on the scalp, midline between the international 10–20 Cz and Fz electrode positions. The scalp data were acquired at a sampling rate of 512 Hz. The effect of ANT stimulation (2 Hz, 7 Hz, and high frequency (>100 Hz)) on HPC iEEG recordings, behavioral state classification based on HPC recordings, and sleep architecture was investigated. The details of the stimulation protocol are shown in Table S1 in the Supplementary Materials.

#### 2.1.3 Sleep Scoring Using Scalp EEG

For visual expert sleep stage scoring, all the PSG recordings were bandpass filtered between 0.3 and 75 Hz with a 60 Hz notch filter using six-order zero-phase Butterworth filters. We used electrodes that were placed in the standard 10–20 system locations. This included eye and chin electrodes for evaluation of eye movements and muscle activity for REM sleep scoring. Visual sleep scoring was done manually by an expert reviewer (EKS) in accordance with guidelines of American Association of Sleep Medicine [40] using a visualisation and analytical software tool CyberPSG (Certicon a.s.). Wakefulness was determined by the presence of eye blinks visualized in frontal scalp and eye leads, accompanied by posteriorly dominant alpha rhythm (8–12 Hz), comprising >50% of the epoch. N2 sleep was scored when low frequency delta activity was present accompanied by K-complexes or spindles. Slow-wave sleep (N3) was scored when high-voltage (>75 μV), delta (0.5 – 4 Hz) activity on scalp EEG was present in at least 20 % (6 sec) of the epoch in the frontal electrode derivations. A similar approach was used in previous studies [22,23,41]. In total, 12,182 sleep epochs were annotated by an expert reviewer using the available PSG recordings. The distribution between individual sleep stages was Awake - 29.4 %, N1 - 4.8 %, N2 37.7 %, N3 - 15.7 % and REM - 12.32 %. The block diagrams for experiment setup and data pipelines are shown in Figure 1.

**Figure 1.**
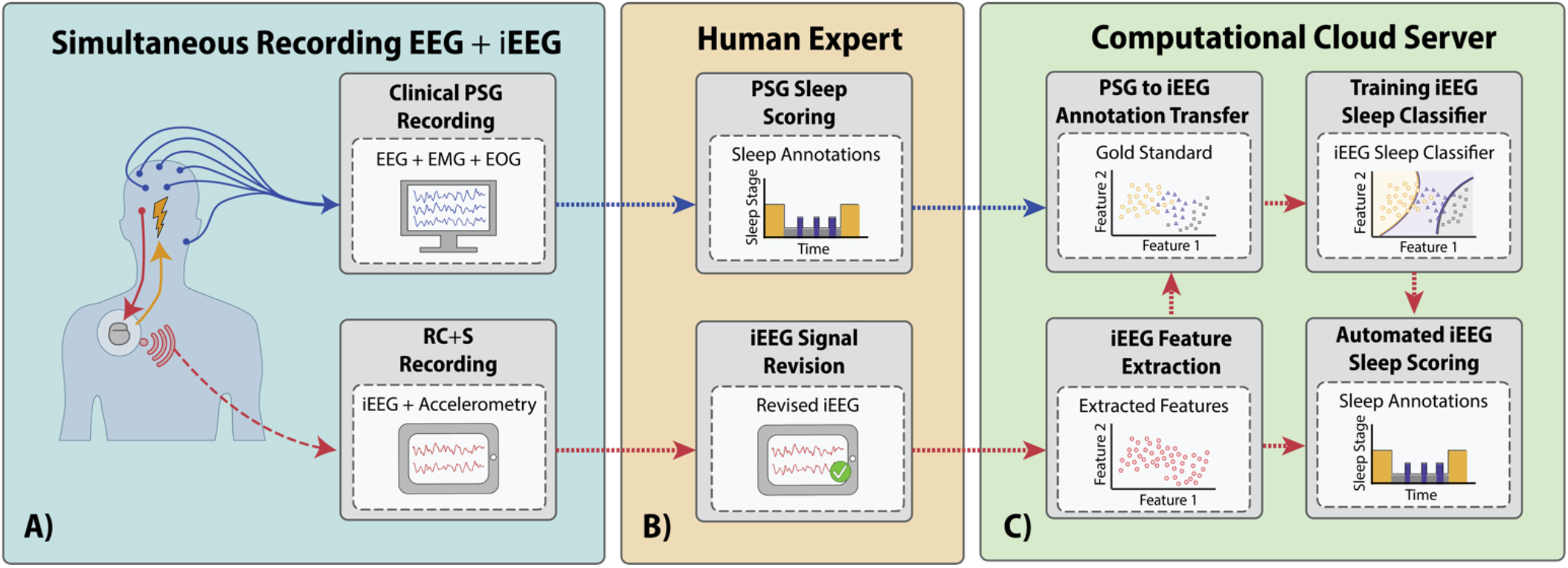
Block-diagram of the process for training, validation, and testing of an automated sleep classifier using simultaneous polysomnography (PSG) and intracranial EEG (iEEG) recordings. ***A)*** *Schematic of the simultaneous scalp and iEEG recordings* ***B)*** *The expert gold standard sleep scoring from PSG is used to create labeled data to develop the automated sleep classifier* [38]. The iEEG was reviewed and the channel with the lowest epileptiform activity and stimulation related artifacts was selected ***C)*** *Direct comparison of the expert sleep scores are used to evaluate performance of the automated iEEG based classifier*.

#### 2.1.4 Selection of an Intracranial Electrode for Sleep Scoring

The investigational Medtronic Summit RC+S™ enables iEEG data streaming from a total of 4 bipolar sensing channels at a time, that can be selected from the 16 total electrode contacts. The fact that some electrode contacts are placed in the epileptic focus, a region with a propensity to show pathological activity such as interictal epileptiform discharges (IEDs), led us to investigate models for behavioral state classification using a single bipolar iEEG channel. Our approach uses a single selectable sensing channel that has a minimum of pathological activity and is optimal for sleep scoring. Individual channels were evaluated for each subject based on the following factors: rate of IEDs, rate of subclinical seizures, and magnitude of DBS artifacts. For the automated classifier we select the HPC electrode with the fewest of all the above.

The impact of ANT stimulation on HPC iEEG signal spectrum for cases of 2 and 7 Hz low frequency stimulation shows that the power spectrum is disrupted with peaks in bands of stimulation frequency and its harmonics (Figure 2). Within the PIB features used in this study the high frequency stimulation (HF>100 Hz) has little impact. To illustrate the variable IED-rate within a single subject, patient H1 exhibited 500 - 5,200 IEDs per hour across different recording channels (Figure 3) and behavioral states [38]. DBS-induced artifacts are also an important factor to consider during electrode selection.

**Figure 2.**
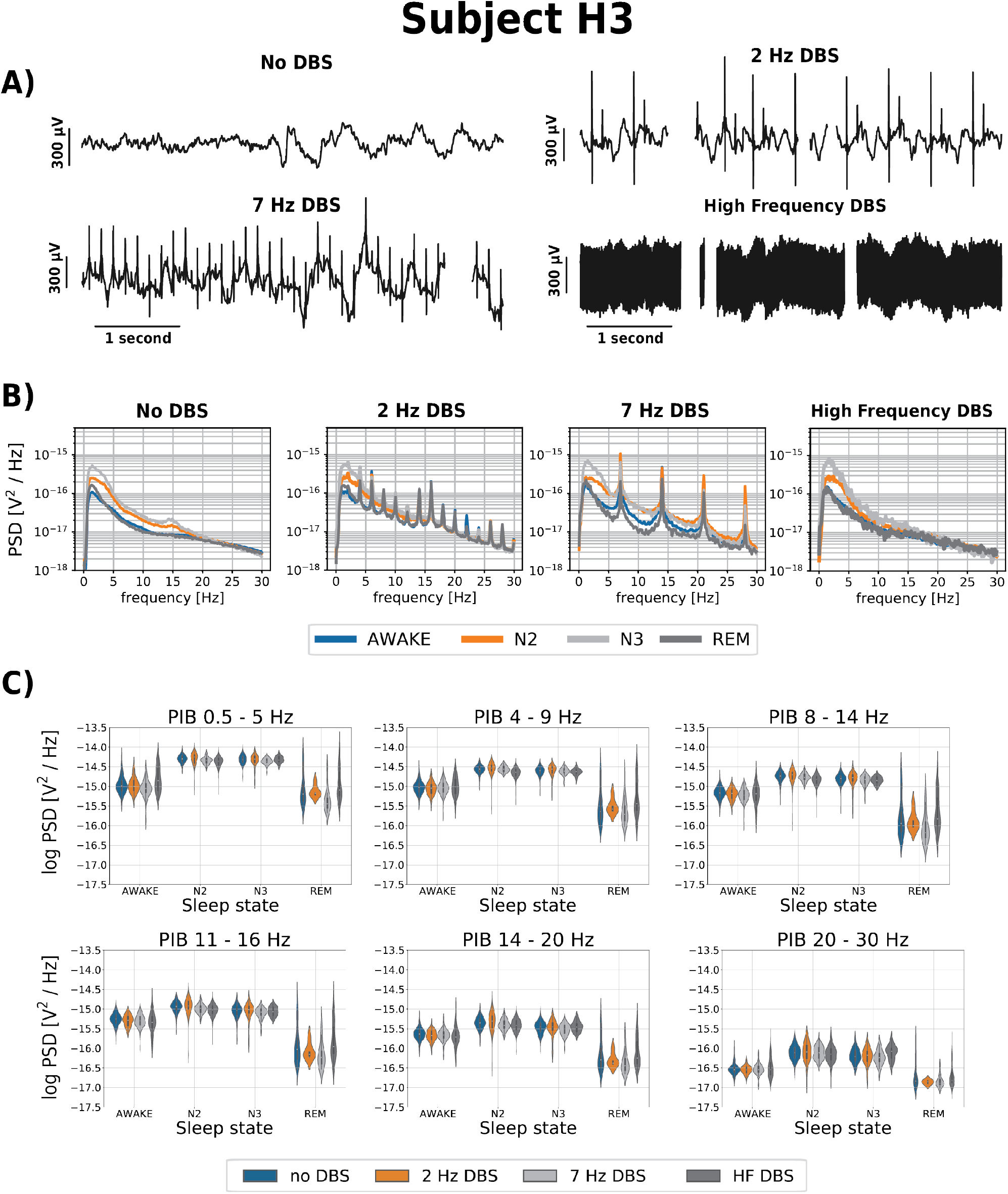
The effect of ANT DBS on iEEG signals. Representative data from subject H3. **A)** The iEEG recorded from right hippocampus (HPC) for different ANT DBS frequencies (No DBS, 2, 7 Hz and high frequency (HF >100 Hz) DBS). The ANT stimulation artifact is clearly apparent on the HPC iEEG timeseries and at high frequency stimulation obscures the iEEG signal **B)** The power spectral density (PSD) for different ANT DBS frequencies (No DBS, 2 Hz, 7 Hz and HF DBS) show DBS artifacts. The figure shows the average of each sleep phase spectrum across three nights using 30-second window, estimated by Welch’s method (Awake, N2, N3, REM). **C)** Power in band features (0.5 - 5 Hz; 4 – 9 Hz; 8 – 14 Hz; 11 – 16 Hz, 14 – 20 Hz, 20 – 30 Hz) extracted from raw HPC iEEG signals from each behavioral state (Awake, N2, N3, REM), over three nights using 30-second window. These data from subject H3 are representative of all patients (see supplementary data).

**Figure 3.**
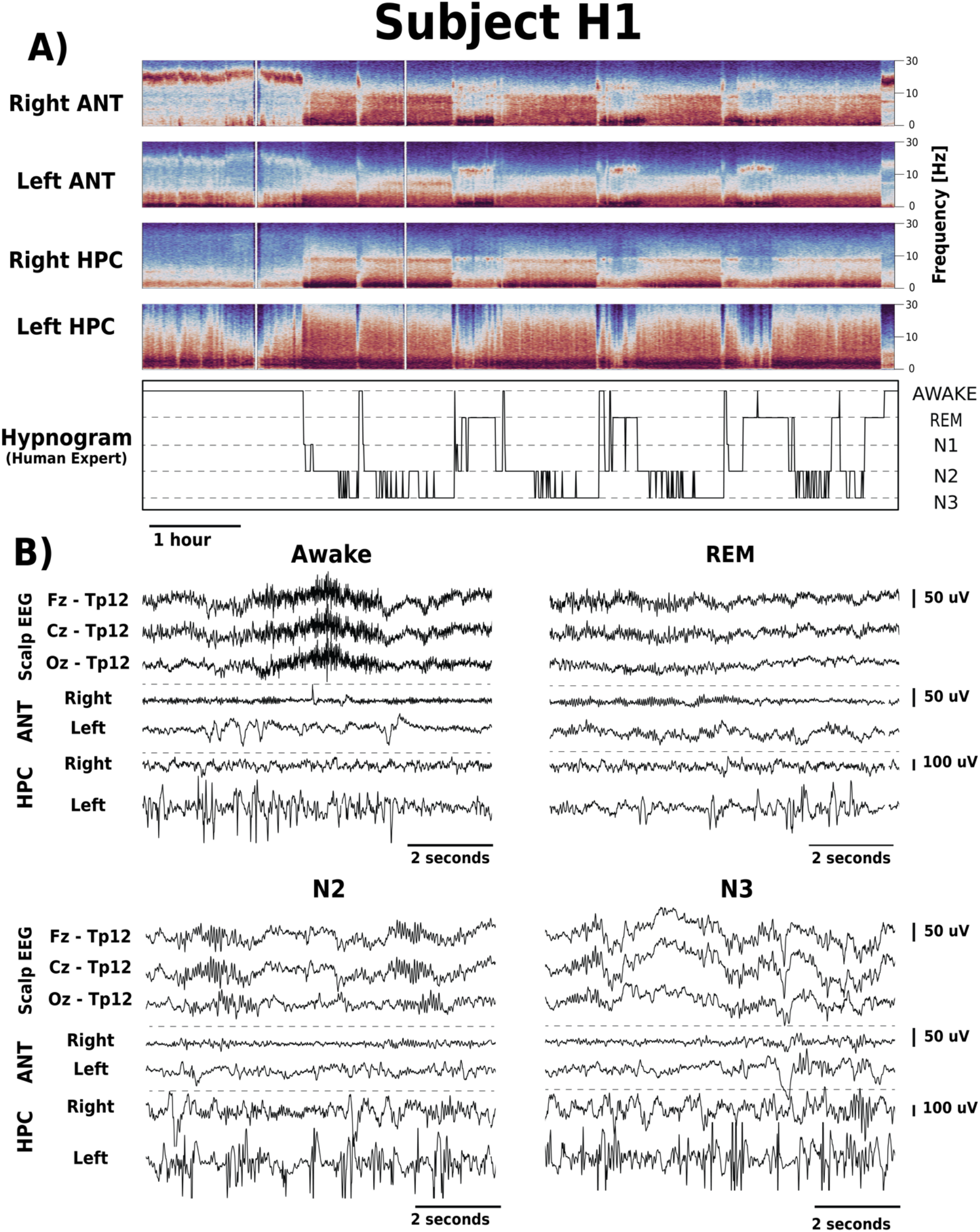
Intracranial EEG (iEEG) signal changes between different behavioral states (Awake, REM, N2 and N3). **A)** Spectrograms of iEEG signals recorded from right and left anterior nucleus of thalamus (ANT) and hippocampus (HPC), show Awake, REM, and non-REM changes. There are differences between iEEG signals recorded from right and left ANT and HPC related to the electrophysiological signatures of epilepsy e.g., interictal epileptiform discharges are increased in the left HPC (patient H1). **B)** Simultaneous scalp-EEG (Fz, Cz and Oz referenced to TP12) and iEEG (bipolar Left ANT, Right ANT, Left HPC, Right HPC) recordings for Awake, REM, N2 and N3.

### 2.2 Automated Sleep Classification

We implemented a classification model utilizing a Naïve Bayes Classifier [35,36,42] with relative power in band (PIB) features extracted from a single iEEG channel to classify 30 second long epochs into the standard sleep categories Awake, N2, N3, REM. The sleep state N1 was excluded due to the insufficient number of samples among the available gold standard data (Table 1).

**Table 1.**
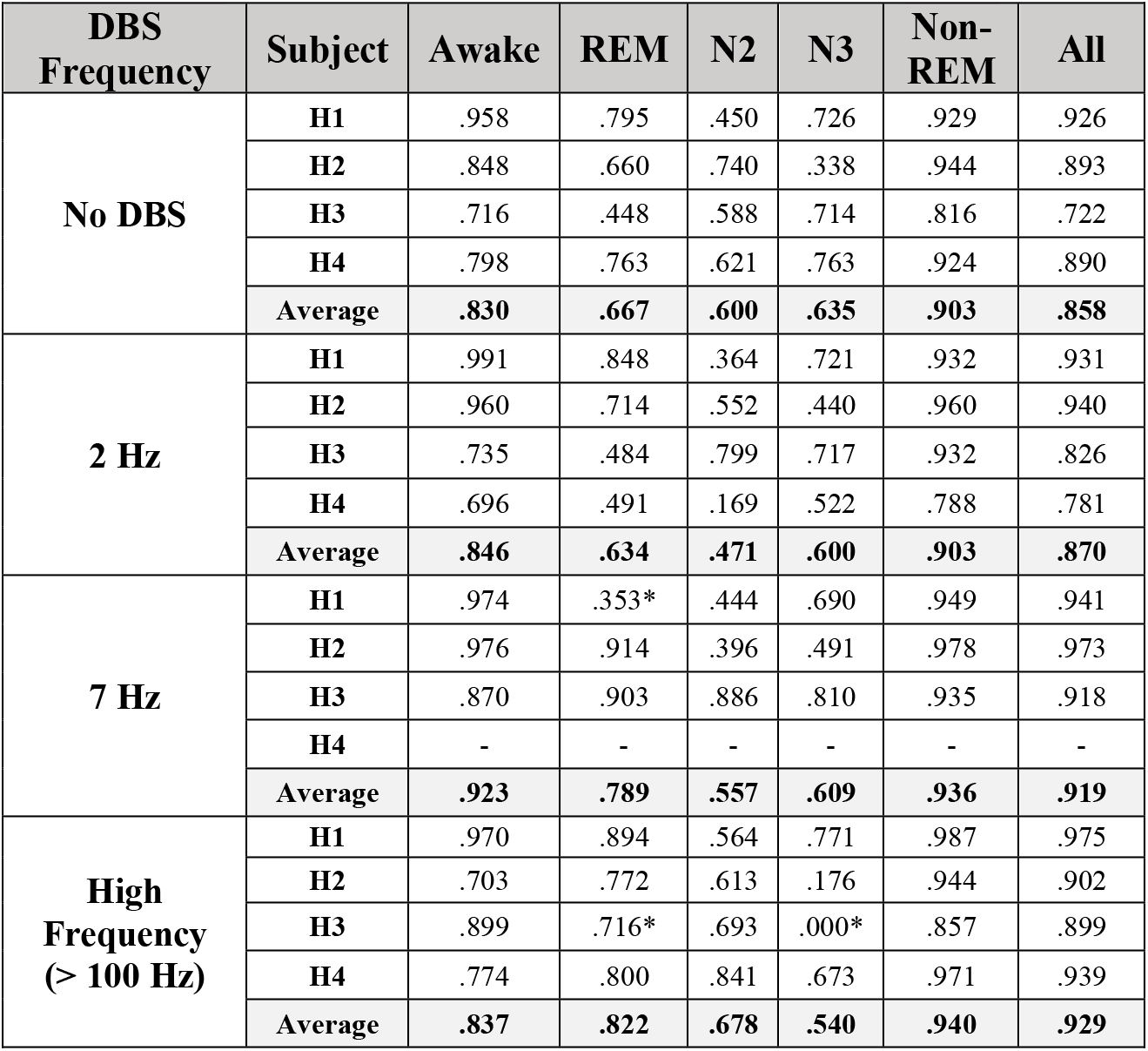
Results of automated behavioral state classification (F1-score) into Awake, REM, non-REM (N2, N3) sleep categories for all subjects under various settings of electrical deep brain stimulation (DBS) in anterior nucleus of thalamus (ANT). Values marked by * were achieved using fewer than 10 samples and are considered less reliable, however, stated for consistency.

| DBS Frequency | Subject | Awake | REM | N2 | N3 | Non-REM | All |
| --- | --- | --- | --- | --- | --- | --- | --- |
| No DBS | H1 | .958 | .795 | .450 | .726 | .929 | .926 |
|  | H2 | .848 | .660 | .740 | .338 | .944 | .893 |
|  | H3 | .716 | .448 | .588 | .714 | .816 | .722 |
|  | H4 | .798 | .763 | .621 | .763 | .924 | .890 |
|  | Average | **.830** | **.667** | **.600** | **.635** | **.903** | **.858** |
| 2 Hz | H1 | .991 | .848 | .364 | .721 | .932 | .931 |
|  | H2 | .960 | .714 | .552 | .440 | .960 | .940 |
|  | H3 | .735 | .484 | .799 | .717 | .932 | .826 |
|  | H4 | .696 | .491 | .169 | .522 | .788 | .781 |
|  | Average | **.846** | **.634** | **.471** | **.600** | **.903** | **.870** |
| 7 Hz | H1 | .974 | .353* | .444 | .690 | .949 | .941 |
|  | H2 | .976 | .914 | .396 | .491 | .978 | .973 |
|  | H3 | .870 | .903 | .886 | .810 | .935 | .918 |
|  | H4 | - | - | - | - | - | - |
|  | Average | **.923** | **.789** | **.557** | **.609** | **.936** | **.919** |
| High Frequency (> 100 Hz) | H1 | .970 | .894 | .564 | .771 | .987 | .975 |
|  | H2 | .703 | .772 | .613 | .176 | .944 | .902 |
|  | H3 | .899 | .716* | .693 | .000* | .857 | .899 |
|  | H4 | .774 | .800 | .841 | .673 | .971 | .939 |
|  | Average | **.837** | **.822** | **.678** | **.540** | **.940** | **.929** |

#### 2.2.1 Data Processing

The iEEG and PSG recordings were stored in the Multi-Scale Electrophysiology Format (MEF) [43] and imported into Python using a Python library “*pymef”* available on GitHub https://github.com/msel-source/pymef. The sleep scoring gold standard annotations were exported from the CyberPSG software and imported into Python using software package *“Python Invasive Electrophysiology Signal Processing Toolbox”* (PiesPro).

The iEEG signals used for automated sleep scoring were filtered using 3rd order IIR Butterworth zero-phase filter with a 40 Hz cutoff frequency. Subsequently, the iEEG signals were segmented into 30-second-long epochs with gold standard annotations. All epochs with more than 15% of samples missing due to packet drops in the wireless data iEEG streaming were excluded, leaving a total of 11,091 epochs from all subjects. The segments were distributed across all sleep stages (Awake – 35 %; N 1 - 5%; N 2 – 34%; N 3 – 15%; REM – 11 %), and across different stimulation frequencies (no-DBS – 52%; 2 Hz DBS – 34 %; 7 Hz DBS – 7 %; high-frequency DBS - 22 %). Additional details are provided in Table S2 in the Supplementary Materials.

Missing iEEG samples within short packet drops were replaced by the average value of the corresponding 30-second epoch. The iEEG data were then filtered by a 0.5-40 Hz band-pass filter and power spectral density (PSD) estimated using Welch’s method for each 30-second-long epoch using a 10-second window with 5 second overlap.

We extracted two sets of PIB features from the following bands: 0.5-5 Hz, 4-9Hz, 8-14Hz, 11-16Hz, 14-20Hz (low beta) and 20-30Hz (high beta) for each 30-second epoch corresponding to the gold standard sleep score. Frequency bands were selected based on prior automated sleep classification work using intracranial and scalp EEG [22–25,37,38,40] and preliminary analysis of patient H1. The first set of features was PIB relative to the power of the whole spectrum from 0.5 – 30 Hz. The second set of features consisted of the relative PIB calculated as a ratio of frequency bands. The PIB ratio was estimated for all non-repeating two-sample ascending combinations within the set of all frequency bands. Subsequently, a decadic logarithm transformation was applied to all features introduced above.

#### 2.2.2 Cancelling of Band Power at Stimulation Frequencies

DBS-induced artifacts alter the PIB features as evident in Figure 2. To reduce the impact of DBS-induced artifacts on extracted features, and thus on classification performance, we applied PIB cancelling - setting to zero the PIBs most effected by DBS artifact. In this way, we removed frequency bands significantly impacted by the DBS-induced stimulation artifacts – 2 & 7 Hz and higher harmonic frequencies. Based on a preliminary data inspection, we eliminated the frequency band 1.5 – 2.5 Hz for 2 Hz stimulation, and the 6 – 8 Hz band for the 7 Hz stimulation and corresponding higher harmonic frequencies. PIB cancelling was performed separately for each stimulation paradigm.

#### 2.2.3 Modes of Operation: Chronic Automated Classification under Different Stimulation Settings

We trained a patient-specific classifier using hippocampal iEEG data during stimulation-free periods then tested and validated the classification performance using data recorded under different ANT stimulation programs (2 Hz, 7 Hz, HF > 100 Hz DBS). There are practical limitations to exposing patients to multiple nights of monitoring in the EMU to screen possible stimulation programs and evaluate the impact of thalamic stimulation on the precision and stability of the automated sleep classifier. We tested the feasibility of transferring classification models (TM) between no DBS and multiple DBS frequencies (2 Hz, 7 Hz, HF (>100 Hz)) separately (Figure 4). Therefore, we validated the classifiers in a scenario when only a single night of concurrent PSG and iEEG is available. We also investigated the impact of PIB cancelling on classification performance by training and validating classifiers for all subjects under different PIB cancelling protocols: no PIB cancelling and DBS frequency specific PIB cancelling (Table S2 Supplementary Materials, Figure 4).

**Figure 4.**
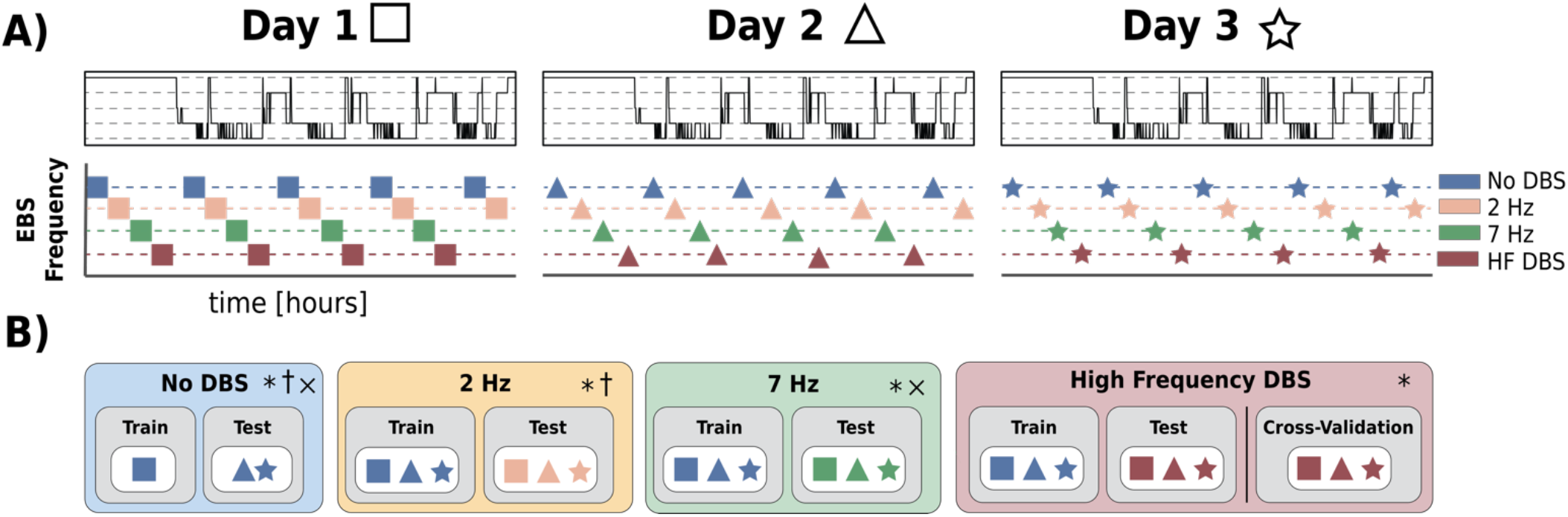
An illustrative scheme of iEEG data utilization throughout the training and testing of classifiers under multiple ANT stimulation programs. The order of data collection for patients varied based on the data collected during the previous nights. **A)** Illustration of iEEG data sampling under different ANT stimulation programs. **B)** A scheme for iEEG data utilization during training and testing of automated sleep classification under different ANT stimulation programs (No DBS, 2 Hz, 7 Hz, HF > 100Hz DBS). Each experiment was performed **\*** - without applying the method of PIB cancelling; **†** - with PIB cancelling specific to 2 Hz stimulation; **×** - with PIB cancelling specific to 7 Hz stimulation. Details are listed in *Table S1 and Table S2 in Supplementary Materials*.

##### a) No DBS

In the first experiment, we used iEEG data collected from three consecutive days of HPC iEEG recording (H1-H4) in the absence of ANT DBS to classify Awake, REM, and non-REM (N2 & N3) sleep stages. Expert sleep scoring annotations with and without ANT DBS were created for all patients. We used available iEEG data acquired without DBS during the first night to train the classifier and data acquired during the second and third night for pseudo-prospective testing. The nights without DBS, however, were not consistent across patients. If data without DBS were not available for the first night, the data without DBS acquired during the second night were used for training and the third night data were utilized for testing (Table S2 Supplementary Materials). This experiment was performed for all patients (H1-4).

##### b) Low Frequency DBS

We investigated the feasibility of deploying the classifiers trained using iEEG data recorded without DBS present to cases with low frequency (2 & 7 Hz) ANT DBS and contaminated by DBS-induced artifacts. The experiment was performed with and without the stimulation active PIB cancelling for all subjects.

##### c) High Frequency DBS

Like the previous experiment, we tested the feasibility of reusing the stimulation-free classifier for the iEEG data recorded while delivering high frequency (>100 Hz) thalamic stimulation.

Additionally, we performed a leave-out cross validation (CV) testing for high frequency DBS data in each patient since we hypothesize that high frequency DBS may decrease signal-to-noise ratio and alter HPC iEEG beyond simple stimulation artifacts. The CV testing was performed in 100 iterations, with 80% of all iEEG data recorded during high frequency DBS. The rest of the data was used for testing. The data were sampled proportionally in all classification categories.

#### 2.2.4 Classification – Naïve Bayes

We developed a classification model utilizing a Naïve Bayes Classifier [35,36,42]. The model uses PIB features extracted in 30-second epochs as an input. The Naïve Bayes classification model is based on Bayes’ Theorem, predicting a conditional probability *P*(*S*|*X*) of the event S (S is a behavioral state {Awake, N2, N3, REM}) given the feature vector X. *P*(*X* |*S*) is obtained as a Probability Density Function of the extracted feature vector for each behavioral state S. The statistical distribution for each of the behavioral states is estimated using training data. *P*(*S*) is a prior probability of a state S and *P*(∅*S*) is defined as 1 − *P*(*S*). For this work, we used a kernel of a multivariate normal distribution with prior probability evenly distributed across all classification classes.

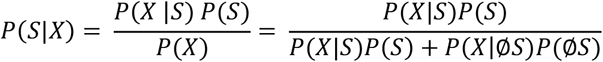

#### 2.2.5 Post-Processing: Hypnogram Correction Rules for Automated Long-Term Data with Dropouts

In this study the encrypted iEEG data are wirelessly transmitted between different hardware devices (implanted neural stimulator, tablet, and cloud) [28,39]. Wireless data transmission in the ambulatory environment can suffer from data loss in the form of dropped data packets due to poor wireless signal connections between individual devices. Both short (< 5 minutes) and long (> 5 minutes) data drops were present in our recordings. Short data drops account for up to 2.9 hours of missing data every day (patients H1-H4). To address the issue of missing data during different behavioral states and transitions we employed a heuristic approach to assign short data drops to specific classes:

1. If a short data drop (< 5 minutes) is preceded by at least 5 minutes of a classified sleep stage that consistently yielded the same sleep stage, the data drop is assigned the sleep stage of the preceding prediction.
2. If a short data drop (< 5 minutes) is preceded by at least 5 minutes of “Awake” state predictions and followed by a REM stage prediction, the data drop is assigned to “Awake”.
3. Any epoch that is classified as “REM” sleep that follows at least 5 minutes of “Awake” is assigned to “Awake”.
4. If a sequence of predicted sleep states with a duration of at least 5 minutes consists of any combination of “N2” and “N3” and is followed by a data drop with a maximum duration of 5 mins, and the following predicted sleep state is either “N2” or “N3”, the void is assigned to a generic non-REM sleep category “N”.
5. If a sequence of predicted sleep states with a duration of at least 5 minutes consists of any combination of “N2”, “N3” and “REM” and is followed by a data drop with a maximum duration of 5 mins, and the following predicted sleep state is either “N2”, “N3” or “REM”, the void is assigned to a generic sleep category “SLEEP”.

### 2.3 Ambulatory Behavioral State Classification in Ambulatory Subjects in Naturalistic Environment

The trained models for automated iEEG behavioral state classification were deployed on a handheld epilepsy personal assist device (EPAD) and integrated with a cloud infrastructure for automated behavioral state classification [38, 39]. Over 190 days was analyzed in four people with epilepsy (H1-4).

### 2.4 Statistics

Remaining cognizant of the unbalanced datasets in this study, we calculated the F1-score classification metric for binary classification of each category and weighted the F1-score average for multi-class classification. The F1-score was calculated as follows

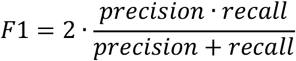

Precision, or positive predictive 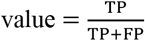 and recall, or 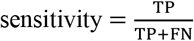 are defined in terms of true positive (TP), false positive (FP) and false negative (FN) classification.

### 2.5 Reproducible Research – Code Sharing

The Bioelectronics Neurophysiology and Engineering Lab is committed to sharing data and code to facilitate reproducible research. All codes are publicly available on Github in a Python software package called *“Python Invasive Electrophysiology Signal Processing Toolbox”* (PiesPro) available on https://github.com/mselair/PiesPro. The data are available upon request.

## 3. Results and Discussion

We developed and tested an approach for fully automated behavioral state classification using a single bipolar iEEG channel recorded from HPC in patients with epilepsy implanted with the investigational Medtronic Summit RC+S™ system.

We recorded continuous iEEG and PSG data in the hospital epilepsy monitoring unit for three consecutive days and nights in four patients. We trained, validated, and tested the automated behavioral state classification using visually scored, gold standard polysomnography. We investigated and verified that the classifier can be trained on data from one night and deployed under various ANT DBS paradigms (high and low frequency stimulation) with high accuracy classification scores. Thereby, we demonstrated for the first time that automated sleep classification based on a single HPC iEEG channel is feasible during concurrent low- or high-frequency ANT DBS. We then deployed the classifiers onto a patient handheld tablet device [39] to track longitudinal sleep patterns in four patients with epilepsy living in their natural environment.

### 3.1 Automated Sleep Scoring Using iEEG Recordings without Concurrent Electrical Deep Brain Stimulation

In the absence of ANT stimulation, the overall classification performance was 0.858 (Table 1) and the results suggest that classifying Awake/non-REM/REM states from a single hippocampal sensing channel has good precision and sensitivity for each behavioral state, with the worst performance for REM sleep with an average F1-score of 0.667. While classification of combined N2 & N3 as non-REM slow wave sleep is very reliable with F1-score 0.940, classifier performance for N2 and N3 individually, is modest HPC (F1_N2_ = 0.600, F1_N3_=0.635). Detailed results for all subjects are shown in Table 1. The features that distinguish N2 from N3 sleep may be difficult to capture in our classification approach which relies on 30 second power in band features. We speculate that more prominent IED activity in HPC [38] in both N2 and N3 sleep stages might drown out the relatively subtle difference of N2 and N3 in PIB features extracted from iEEG signals and create larger classification errors between N2 and N3.

We validated the feasibility of automated iEEG sleep classification using a single HPC iEEG. Previous studies have investigated classification of long-term iEEG into awake and non-REM sleep (N2, N3) [22, 23], but REM sleep classification was not addressed. Chen et al. utilized PSG-based annotations to build an automated sleep classifier that was able to differentiate REM with some accuracy (sensitivity 65.4% and specificity 57.6%) using iEEG from sub-thalamic nucleus in people with Parkinson’s disease [44]. In the future it will be interesting to investigate the iEEG correlates of specific brain structures to better understand the electrophysiology correlates of different behavioral states.

### 3.2 Assessment of Automated Sleep Scoring from Hippocampal iEEG During Low Frequency and High Frequency Electrical Brain Stimulation

We trained and tested a behavioral state classification model for multiple DBS frequencies (2 Hz, 7 Hz, HF > 100Hz) using a single bipolar iEEG channel with available gold standard polysomnography annotations acquired during three consecutive nights for the subjects (H1-4).

ANT stimulation can influence hippocampal background activity and elicit evoked hippocampus responses [45]. Despite the potential impact of ANT stimulation on HPC-based sleep classification, we observed acceptable classifier performance during both high and low frequency ANT stimulation. Although 2 Hz DBS, corrupted the iEEG power spectrum at the stimulation frequency and its harmonics (2, 4, 6Hz, etc), the average performance across all subjects was good (F1_ALL_ = 0.870). Awake (F1_Awake_ = 0.846) and non-REM (_F1non-REM_=0.903) classification precision was superior to REM (F1_REM_ = 0.634). Classifier performance was slightly better during 7 Hz stimulation (F1_ALL_ = 0.919) for Awake (F1_Awake_ = 0.923), and non-REM (F1_non-REM_ = 0.936) states, and notably improved for REM (F1_REM_ = 0.789). The results highlight the challenge of differentiating Awake from REM and N2 from N3 using a single channel of HPC iEEG (Figure 5). We speculate, that the classification challenge between N2 and N3 stages is related to more prominent IED spiking during non-REM sleep and the evoked hippocampal response to low frequency ANT stimulation (2 and 7Hz). The increased IED rate and evoked HPC response influence PIB features and make N2 and N3 PIB less distinct. Lastly, the sharp transient artifact seen during stimulation can elevate high frequency PIB.

**Figure 5.**
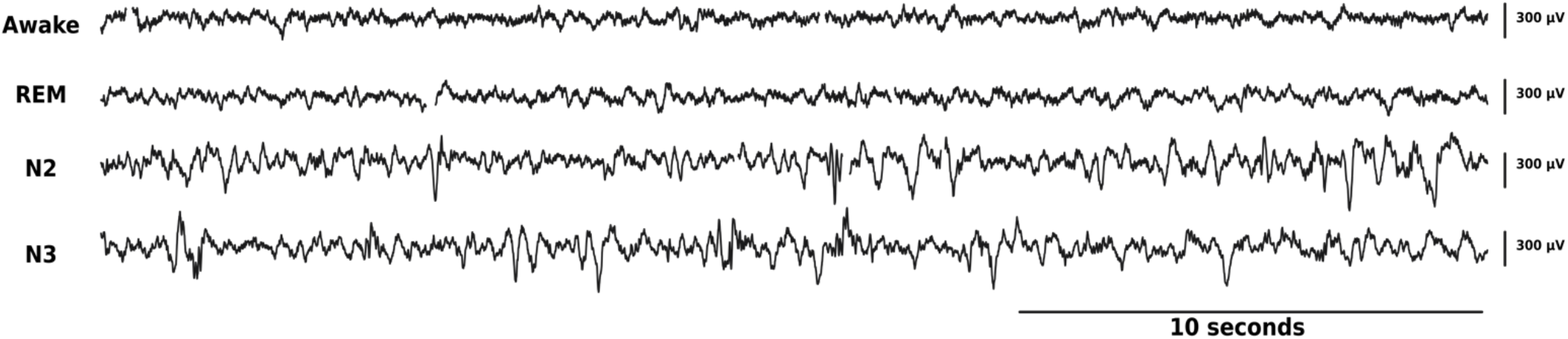
Samples of 30-second epochs of raw data recorded in right HPC channel during multiple behavioral states (gold standard classification): from top to bottom: Awake, REM, N2 and N3.

Given the clinical use of high frequency ANT DBS to treat epilepsy [33], it was important to evaluate the feasibility of re-using a classifier trained on HPC iEEG data without concurrent stimulation to HPC iEEG data acquired during high frequency ANT stimulation. The automated classifier was able to differentiate Awake, non-REM and REM (F1_Awake_ = 0.837, F1_non-REM_ = 940, F1_REM_ = 0.822, and F1_ALL_ = 0.929) behavioral states with good classification scores. The differentiation of N2 from N3, using power in band features as we did here, was modest. Again, the modest N2 and N3 classification might be related to prominent IED spiking activity in non-REM sleep stages. In addition, the neuromodulatory effect of high frequency ANT DBS attenuating the power of iEEG signal in HPC across all frequencies may play a role [45]. Lastly, HF DBS may impact sleep classification performance when stimulation artifacts occur with a frequency close to the Nyquist frequency (half of the sampling frequency) due to an insufficient antialiasing filter. This applies specifically to signals acquired at 250 Hz with concurrent high frequency (>100 Hz) DBS. However, this can be mitigated by increasing the sampling frequency to 500 Hz. Another possible solution for aliasing caused by the presence of high frequency stimulation artifacts is blanking [46,47]. Blanking is a capability of the sensing hardware to disconnect the sensing circuit during electrical stimulation to avoid saturation of the sensing circuit components.

### 3.3 Behavioral State Classification in Ambulatory Subjects in Their Natural Environment

After building the patient-specific models using 3 days of inpatient simultaneous iEEG and PSG recordings, we then prospectively deployed the behavioral state classifiers on continuous, long-term HPC iEEG data totalling over 190 days in four human subjects (H1-H4). The algorithms for automated iEEG behavioral state classification were deployed onto a handheld epilepsy personal assist device (EPAD) and in a cloud system [28,39]. The presentation layer, a web browser interface for long-term iEEG-data (eHealth Dashboard) (Figure 6) monitoring was utilized to present automated sleep staging together with patient seizure and medication logs for physician review (Figure 6). On average, subjects slept 7.91±1.96 hours a day with 5.76±1.23 hours spent in non-REM and 2.05±1.18 hours in REM sleep. Evaluation of the influence of different stimulation paradigms and seizures on sleep architecture remain an interest for future research. The presentation layer providing integration of multiple levels of information (seizure rates, interictal epileptiform discharges, sleep) is essential for this research.

**4.**
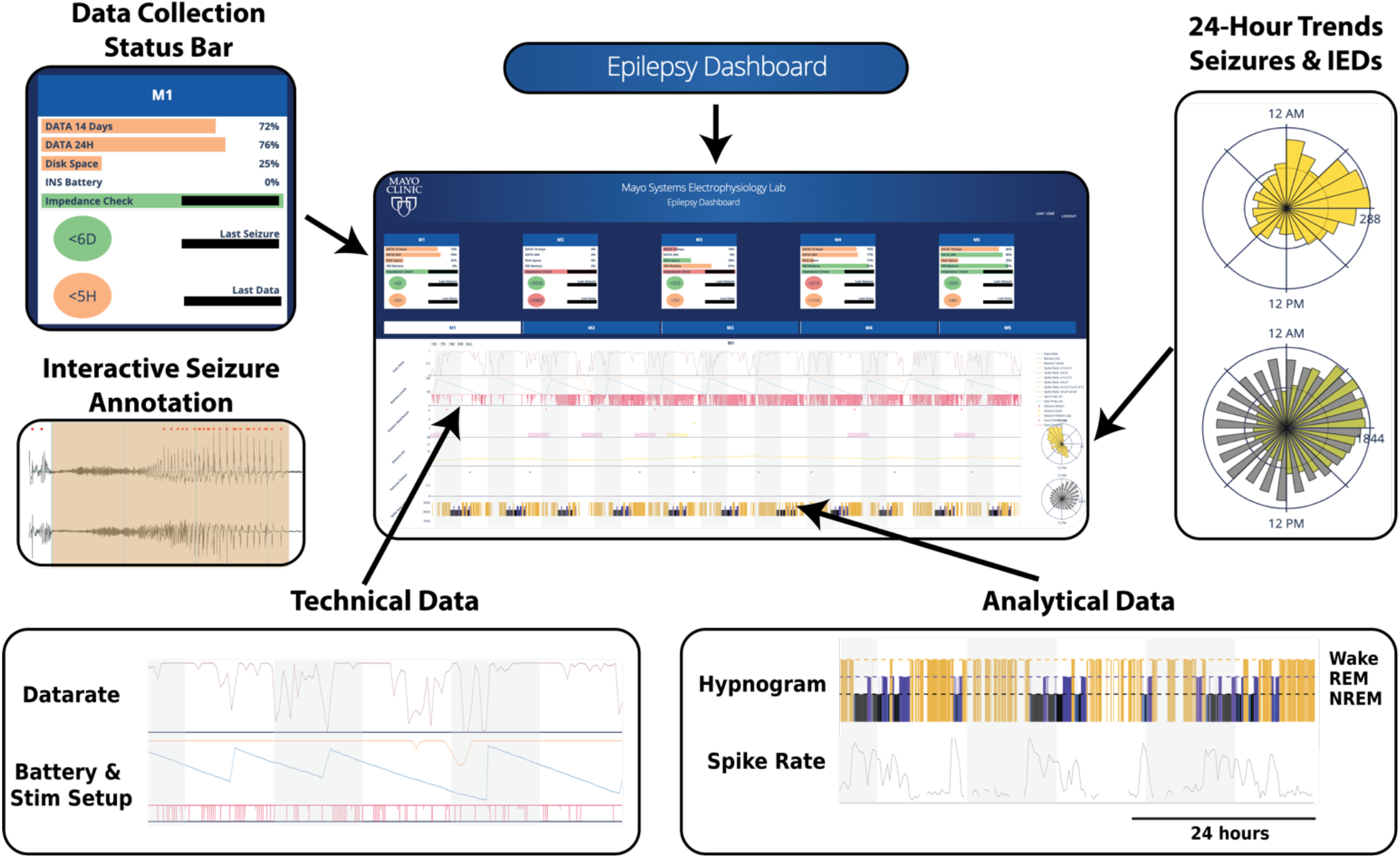
Web-based Epilepsy Dashboard: Trained algorithms deployed on a patient handheld tablet and a cloud system with a web browser user interface for physician to review technical data, patient annotations, gold-standard seizure annotations, and monitoring of long-term iEEG-data, epilepsy biomarkers (such as interictal epileptiform discharge activity), and automated sleep scoring.

### 3.4 Limitations

The main advantage of the proposed method is the ability to continuously track behavioral state using HPC iEEG recordings during therapeutic ANT brain stimulation in ambulatory subjects living in their natural environment. However, there are limitations with respect to classification performance and distinguishing between two non-REM sleep stages, N2 and N3. We speculate that the reason for modest differentiation between N2 and N3 is related to ongoing epileptiform iEEG activity and, possibly the influence of DBS on HPC iEEG activity. Similarly, the differentiation of Awake and REM remains challenging.

The classification of N2 versus N3 slow-wave sleep and REM and Awake state might be improved with additional iEEG channels and other physiologic signals, such as accelerometery, electrocardiogram, and core temperature. Accelerometery data can be streamed by the investigational Medtronic Summit RC+S™ device and is currently under investigation.

Lastly, this study utilizes 3 consecutive nights of simultaneous iEEG and PSG data, but the method was not tested against scalp EEG based PSG outside of the controlled hospital environment. Significant changes in iEEG activity around the time of seizures and with medication changes can impact the electrophysiological data (and related power in bands) and may impair classification. In the future, ambulatory PSG should enable a more comprehensive investigation of long-term behavioral state changes.

## 4. Conclusions

Sleep disruption is a common comorbidity of epilepsy, and an important consideration for DBS optimization. In fact, sleep, cognitive and mood comorbidities have significant impact on quality of life [18]. Therefore, reliable automated behavioral state classification in ambulatory subjects during ANT-DBS is needed to track sleep while optimizing DBS therapy. Currently, the study of sleep disruption in neurologic disease is challenging given the poor correlation between patient reports and gold-standard polysomnograms [20]. The overarching importance of sleep for brain health, and the poor correlation between objective sleep measures and patient reports, has generated enormous interest in non-invasive devices to objectively track sleep in ambulatory subjects. Many non-invasive devices perform poorly [48], but some show good performance when compared to gold-standard polysomnograms [49,50]. While significant advances have been made in ambulatory PSG, with only a few exceptions they are directed at characterizing only a couple of nights of sleep. This does not fill the technology gap for chronic sleep tracking needed for optimal management of neurologic and psychiatric diseases. Patients with implantable neural sensing devices provide a unique platform for long-term sleep investigations and targeting common sleep related comorbidities of neurologic disease. Ambulatory sleep monitoring with implanted devices are now emerging, and have been explored in Parkinson disease [24] and in epilepsy with subscalp sensing [51].

Here we describe an approach for reliable automated behavioral state classification using a single HPC bipolar channel during concurrent ANT DBS. The performance of patient specific behavioral state classification models trained, validated, and tested on concurrently recorded scalp polysomnography and HPC iEEG show good classification of Awake, REM and non-REM (N2+N3) sleep, with and without ANT DBS (2Hz, 7Hz, HF > 100Hz). Automated classification of behavioral states (Awake, non-REM & REM sleep) was deployed on a handheld device integrated with a cloud platform and used to characterize long-term sleep profiles in ambulatory human subjects with epilepsy living in their natural environment. The results also highlight the relative challenge of differentiating Awake from REM and differentiating N2 from N3 using only a single channel of HPC iEEG. The challenge for distinguishing N2 from N3 likely reflects the increase in epileptiform activity in non-REM sleep, the HPC evoked response from ANT DBS, and possibly the closely spaced DBS electrodes that are poorly suited for measuring widespread delta (1 – 4 Hz) activity used for visual sleep scoring with scalp EEG.

This work advances the effort to better understand the bidirectional relationship between sleep, epilepsy, and the impact of DBS. The ability to classify behavioral state with a compact algorithm embedded in an implantable device, or on local and distributed computing resources [21,28,39],enables novel closed-loop stimulation protocols that can adaptively respond to changing brain dynamics. Near term applications include circadian DBS paradigms where stimulation during sleep and awake states are selected to optimize sleep, epilepsy, and comorbidities [52]. The data are mixed as to whether high frequency stimulation commonly used for reducing seizures may negatively impact sleep, mood, and memory [3,19,53]. For patients who commonly have diurnal seizure patterns, such as temporal lobe epilepsy where seizures occur primarily in late morning and early afternoon [13–15], they may benefit from different DBS during the night when the physiological benefits of sleep are critical to normal brain health. Lastly, the selective application of DBS in post-ictal slow wave sleep may prove useful for disrupting pathological seizure related consolidation [41,54].

## Data Availability

Data is available upon reasonable request.

## Funding

This work was supported by NIH Brain Initiative UH2&3 NS095495 *Neurophysiologically-Based Brain State Tracking & Modulation in Focal Epilepsy*, DARPA HR0011-20-2-0028 *Manipulating and Optimizing Brain Rhythms for Enhancement of Sleep (Morpheus)*, Mayo Clinic, and Medtronic Inc. Medtronic provided the investigational Medtronic Summit RC+STM devices. V.K. was partially supported by institutional funding of Czech Technical University in Prague, Czech Republic. Certicon a.s. provided Cyber PSG viewer for research purposes.

## Disclosures

GW, BB, JVG, and BL are named inventor for intellectual property developed at Mayo Clinic and licensed to Cadence Neuroscience Inc. BNL royalties are waived to his Mayo Clinic research account. GW has licensed intellectual property developed at Mayo Clinic to NeuroOne, Inc. BL, GW, and NG in an investigator for the Medtronic Deep Brain Stimulation Therapy for Epilepsy Post-Approval Study (EPAS). VK consults for Certicon a.s., IB has received compensation from an internship with Cadence Neuroscience Inc., for work unrelated to the current publication. Mayo Clinic has received research support and consulting fees on behalf of GW, BNL and BB from UNEEG, NeuroOne Inc., Epiminder, Medtronic Plc., and Philips Neuro.

## Supplementary Materials

### Clinical Information: Neurophysiologically-Based Brain State Tracking & Modulation in Focal Epilepsy (NIH UH3 NS095495 & FDA IDE-G180224)

We consented 6 patients and implanted 4 patients with drug resistant mesial temporal lobe epilepsy (mTLE) as part of the NIH Brain Initiative sponsored *Neurophysiologically-Based Brain State Tracking & Modulation in Focal Epilepsy*. Patients are implanted with investigational Medtronic Summit RC+S™ neural sense and stimulation device with bilateral anterior nucleus of thalamus (ANT) and hippocampus (HPC) electrodes. The epilepsy patient assistant device (EPAD) is an application running on a hand-held device provides integration of wearable and implantable devices [28,39]. The FDA IDE protocol investigates electrical deep brain stimulation (DBS) paradigms, including low frequency (2 & 7 Hz) and high frequency (> 100 Hz) electrical stimulation, seizure detection and forecasting, behavioral state tracking, and adaptive DBS control. Supplementary Figure 1 shows a schematic of the implanted system and co-registration of implanted electrodes.

**Supplementary Figure 1.**
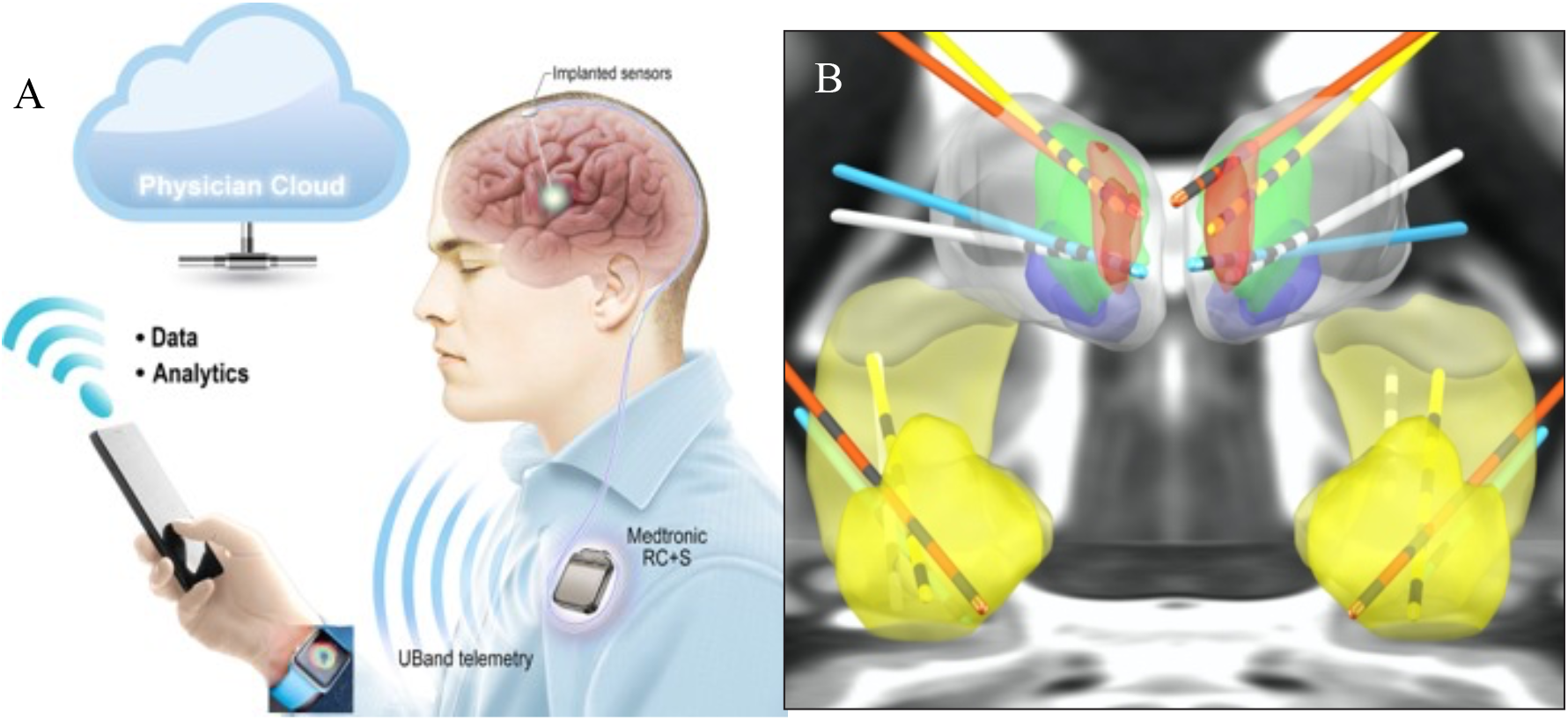
Epilepsy Management System: A) Implantable neural sensing and stimulating (RC+S) device with bi-directional communication to off-the-body computing on EPAD enabling machine learning analytics. The EPAD system integrates RC+S, wearable sensors (e.g., watch), handheld computing device with cloud computing environment. B) Co-registration of the bilateral ANT and HPC leads (4 electrode contacts on each lead) in the four implanted patients. ANT (red) and HPC (light yellow) and AMG (yellow). Epilepsy patient assistance device (EPAD), Anterior nucleus of the thalamus (ANT), Hippocampus (HPC), Amygdala (AMG).

**Supplementary Table S1.** Setup of DBS parameters for subjects during three consecutive nights (Day 1-3) in the hospital epilepsy monitoring unit when the iEEG data were acquired simultaneously with PSG data to create the gold standard human expert sleep scoring used for training, validation, and testing. The iEEG data used for classification were acquired from Hippocampus (HPC). Electrical deep brain stimulation was applied bilaterally in the anterior nucleus of thalamus (ANT). Periodic cycling between different DBS setups, that are given by frequency (f), stimulation current (I) and pulse width (PW), was applied to all subjects.

|  | Day 1 |  |  | Day 2 |  |  | Day 3 |  |  |
| --- | --- | --- | --- | --- | --- | --- | --- | --- | --- |
|  | f (Hz) | I (mA) | PW(us) | f (Hz) | I (mA) | PW(us) | f (Hz) | I (mA) | PW(us) |
| H1 | - | - | - | 7; HF | 3 | 90 | 2; HF | 3 | 90 |
| H2 | 2 | 6 | 200 | 2; 7 | 5; 6 | 200 | HF | 4 | 200 |
| H3 | 2 | 6 | 200 | 7; HF | 4 | 200 | 2; 7 | 5 | 200 |
| H4 | HF | 2 | 200 | 2 | 6 | 200 | 7 | 6 | 200 |

**Supplementary Table S2.** Number of 30-second iEEG epochs collected, DBS parameters, and sleep stage across the three hospital nights. Only epochs with data rate higher than 85% are included. High frequency (HF>100 Hz). Rapid eye-movement (REM) sleep, non-REM (N1, N2, N3) sleep.

|  |  | Night 1 |  |  |  |  | Night 2 |  |  |  |  | Night 3 |  |  |  |  |
| --- | --- | --- | --- | --- | --- | --- | --- | --- | --- | --- | --- | --- | --- | --- | --- | --- |
|  | Stim Mode | W | N1 | N2 | N3 | REM | W | N1 | N2 | N3 | REM | W | N1 | N2 | N3 | REM |
| **H1** | No stim | 421 | 14 | 291 | 266 | 168 | 277 | 6 | 119 | 102 | 91 | 135 | 12 | 75 | 123 | 40 |
|  | 2 Hz | - | - | - | - | - | - | - | - | - | - | 48 | 2 | 34 | 50 | 53 |
|  | 7 Hz | - | - | - | - | - | 148 | 3 | 82 | 30 | 3 | - | - | - | - | - |
|  | HF | - | - | - | - | - | 93 | 5 | 74 | 75 | 38 | 103 | 1 | 46 | 33 | 22 |
|  | All | **421** | **14** | **291** | **266** | **168** | **518** | **14** | **275** | **207** | **132** | **286** | **15** | **155** | **211** | **115** |
| **H2** | No stim | - | - | - | - | - | 320 | 19 | 113 | 63 | 22 | 72 | 7 | 268 | 34 | 54 |
|  | 2 Hz | - | - | - | - | - | 115 | 12 | 49 | 71 | 5 | - | - | - | - | - |
|  | 7 Hz | - | - | - | - | - | 122 | 1 | 69 | 35 | 17 | - | - | - | - | - |
|  | HF | - | - | - | - | - | - | - | - | - | - | 89 | 32 | 347 | 33 | 60 |
|  | All | - | - | - | - | - | **557** | **32** | **231** | **169** | **44** | **161** | **39** | **615** | **67** | **114** |
| **H3** | No stim | 447 | 16 | 296 | 83 | 180 | 171 | 8 | 46 | 25 | 25 | 327 | 14 | 98 | 42 | - |
|  | 2 Hz | 145 | 16 | 282 | 107 | 117 | - | - | - | - | - | 45 | 14 | 58 | 26 | - |
|  | 7 Hz | - | - | - | - | - | 104 | - | 24 | 24 | 23 | 30 | 6 | 80 | 6 | - |
|  | HF Hz | - | - | - | - | - | 100 | 1 | 25 | 8 | 10 | - | - | - | - | - |
|  | All | **592** | **32** | **578** | **190** | **297** | **375** | **9** | **115** | **57** | **58** | **402** | **34** | **236** | **74** | **-** |
| **H4** | No stim | 71 | 42 | 183 | 62 | 59 | 90 | 67 | 220 | 40 | 35 | - | - | - | - | - |
|  | 2 Hz | - | - | - | - | - | 142 | 144 | 427 | 72 | 43 | - | - | - | - | - |
|  | 7 Hz | - | - | - | - | - | - | - | - | - | - | - | - | - | - | - |
|  | HF Hz | 111 | 76 | 370 | 120 | 86 | - | - | - | - | - | - | - | - | - | - |
|  | All | **182** | **118** | **553** | **182** | **145** | **232** | **211** | **647** | **112** | **78** | **-** | **-** | **-** | **-** | **-** |

**Supplementary Figure 2.**
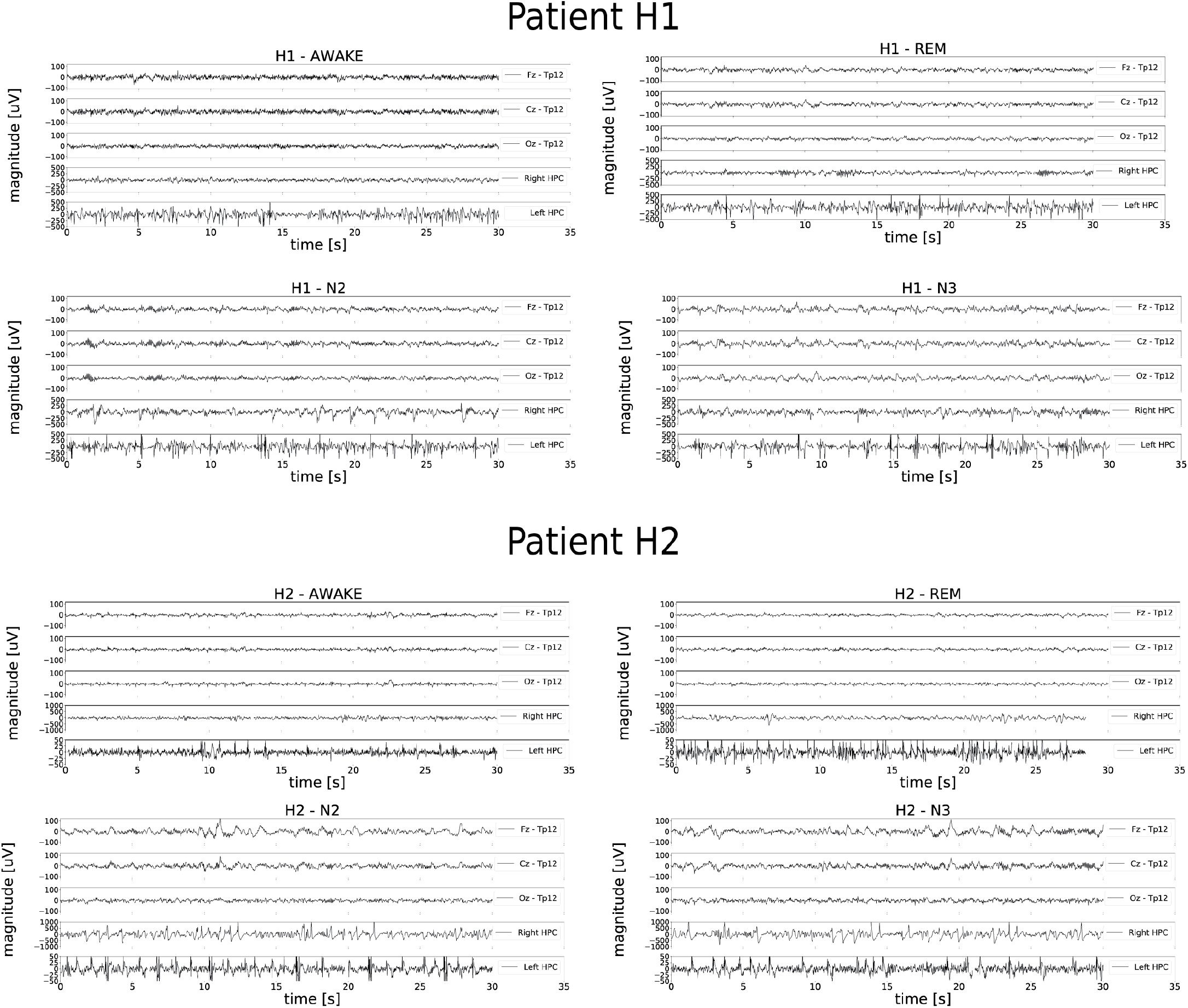
Representative examples of intracranial EEG (iEEG) signal changes between different behavioral states (Awake, REM, N2 and N3) for patients H1 and H2. Simultaneous scalp-EEG (Fz, Cz and Oz referenced to TP12) and iEEG (bipolar Left ANT, Right ANT, Left HPC, Right HPC) recordings for Awake, rapid eye-movement (REM), non-REM (N2 and N3) sleep.

**Supplementary Figure 3.**
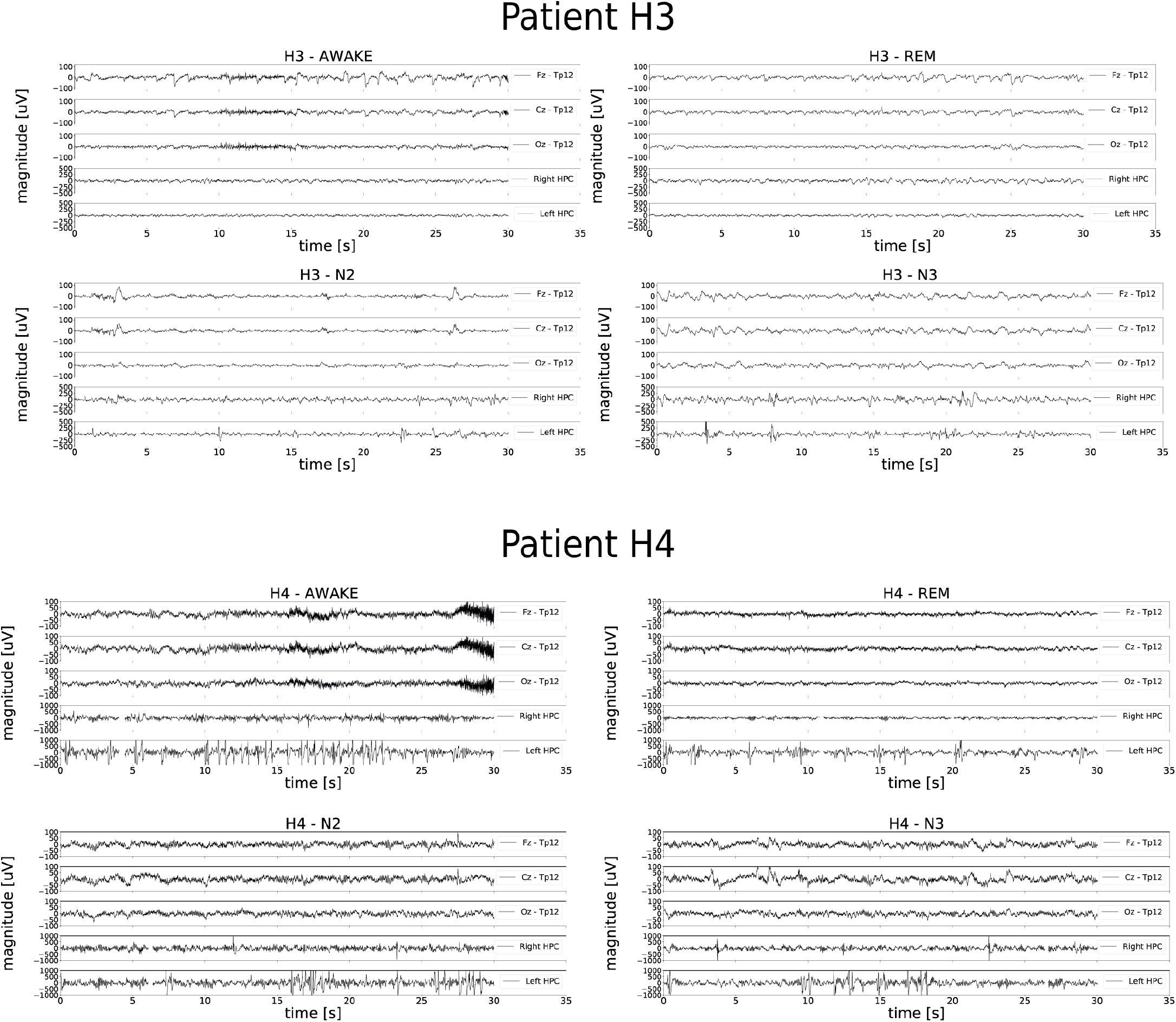
Representative examples of intracranial EEG (iEEG) signal changes between different behavioral states (Awake, REM, N2 and N3) for patients H3 and H4. Simultaneous scalp-EEG (Fz, Cz and Oz referenced to TP12) and iEEG (bipolar Left ANT, Right ANT, Left HPC, Right HPC) recordings for Awake, rapid eye-movement (REM), non-REM (N2 and N3) sleep.

**Supplementary Figure 4.**
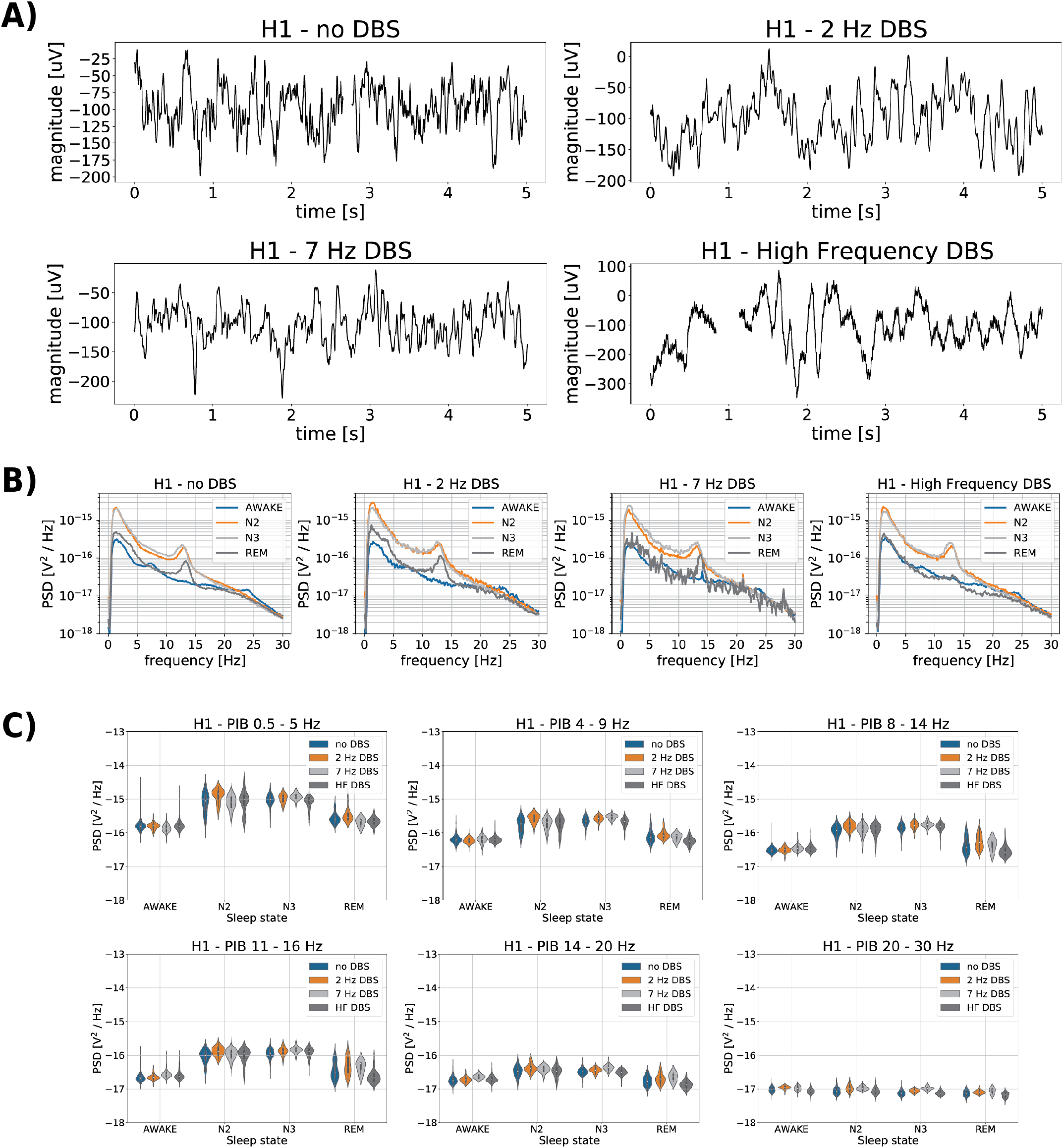
The effect of ANT DBS on iEEG signals. **A)** The iEEG recorded from right hippocampus (HPC) (patient H1) for different ANT DBS frequencies (No DBS, 2, 7 Hz and HF DBS). **B)** Power spectral density (PSD) for different ANT DBS frequencies (No DBS, 2 Hz, 7 Hz and HF DBS): average of each sleep phase spectrum across three nights using 30-second window, estimated by Welch’s method (Awake, N2, N3, REM); **C)** Power in band features (0.5 - 5 Hz; 4 – 9 Hz; 8 – 14 Hz; 11 – 16 Hz, 14 – 20 Hz, 20 – 30 Hz) extracted from raw HPC iEEG signals from each sleep states (Awake, N2, N3, REM), over three nights using 30-second window. These data from subject H3 are representative of all patients. High frequency (HF>100 Hz). Rapid eye-movement (REM) sleep, non-REM (N2, N3) sleep, Anterior nucleus of thalamus (ANT), intracranial EEG (iEEG), Hippocampus (HPC), Deep brain stimulation (DBS), Power spectral density (PSD)

**Supplementary Figure 5.**
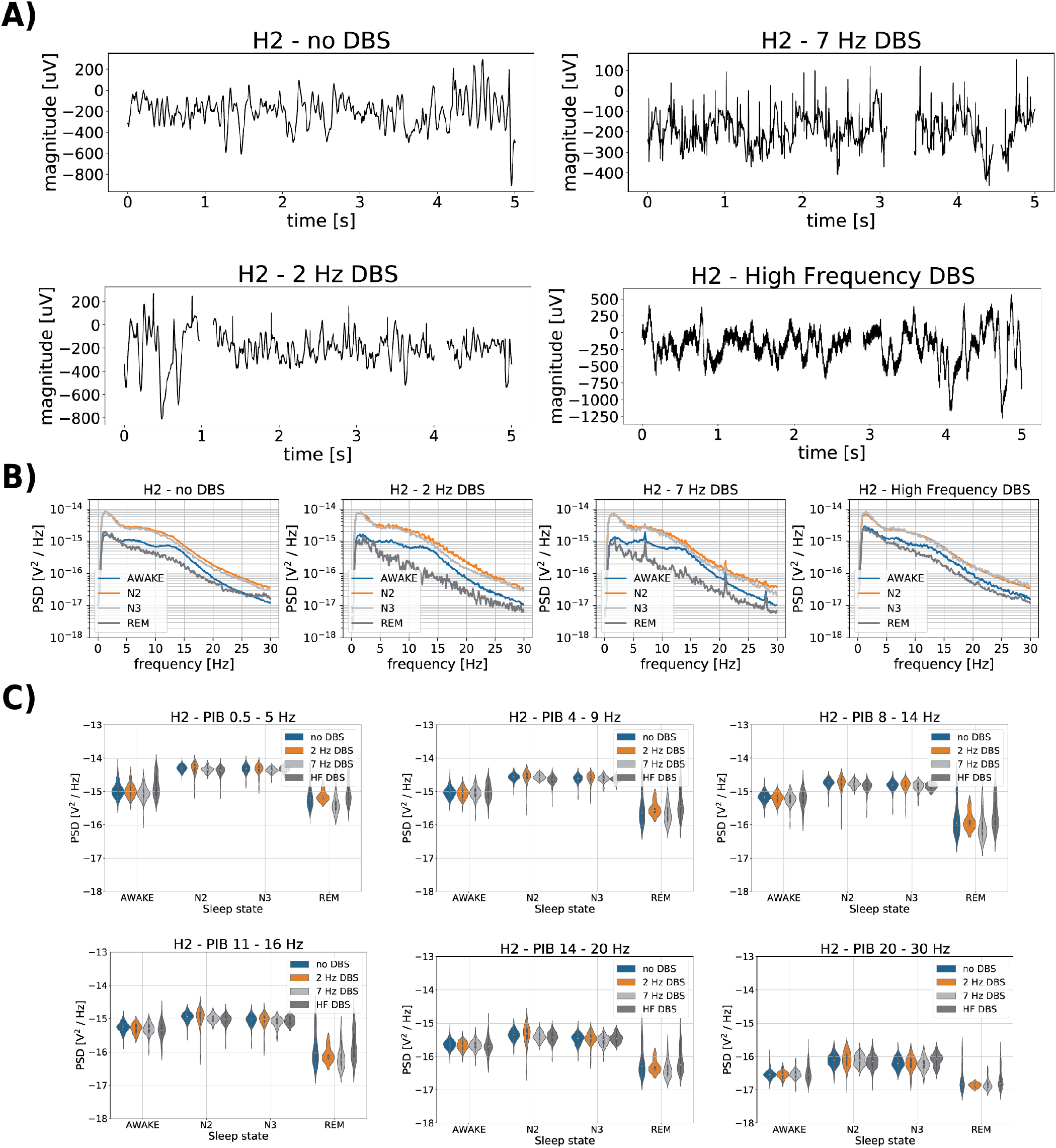
The effect of ANT DBS on iEEG signals. **A)** The iEEG recorded from right hippocampus (HPC) (patient H2) for different ANT DBS frequencies (No DBS, 2, 7 Hz and HF DBS). **B)** Power spectral density (PSD) for different ANT DBS frequencies (No DBS, 2 Hz, 7 Hz and HF DBS): average of each sleep phase spectrum across three nights using 30-second window, estimated by Welch’s method (Awake, N2, N3, REM); **C)** Power in band features (0.5 - 5 Hz; 4 – 9 Hz; 8 – 14 Hz; 11 – 16 Hz, 14 – 20 Hz, 20 – 30 Hz) extracted from raw HPC iEEG signals from each sleep states (Awake, N2, N3, REM), over three nights using 30-second window. These data from subject H3 are representative of all patients. High frequency (HF>100 Hz). Rapid eye-movement (REM) sleep, non-REM (N2, N3) sleep, Anterior nucleus of thalamus (ANT), intracranial EEG (iEEG), Hippocampus (HPC), Deep brain stimulation (DBS), Power spectral density (PSD)

**Supplementary Figure 6.**
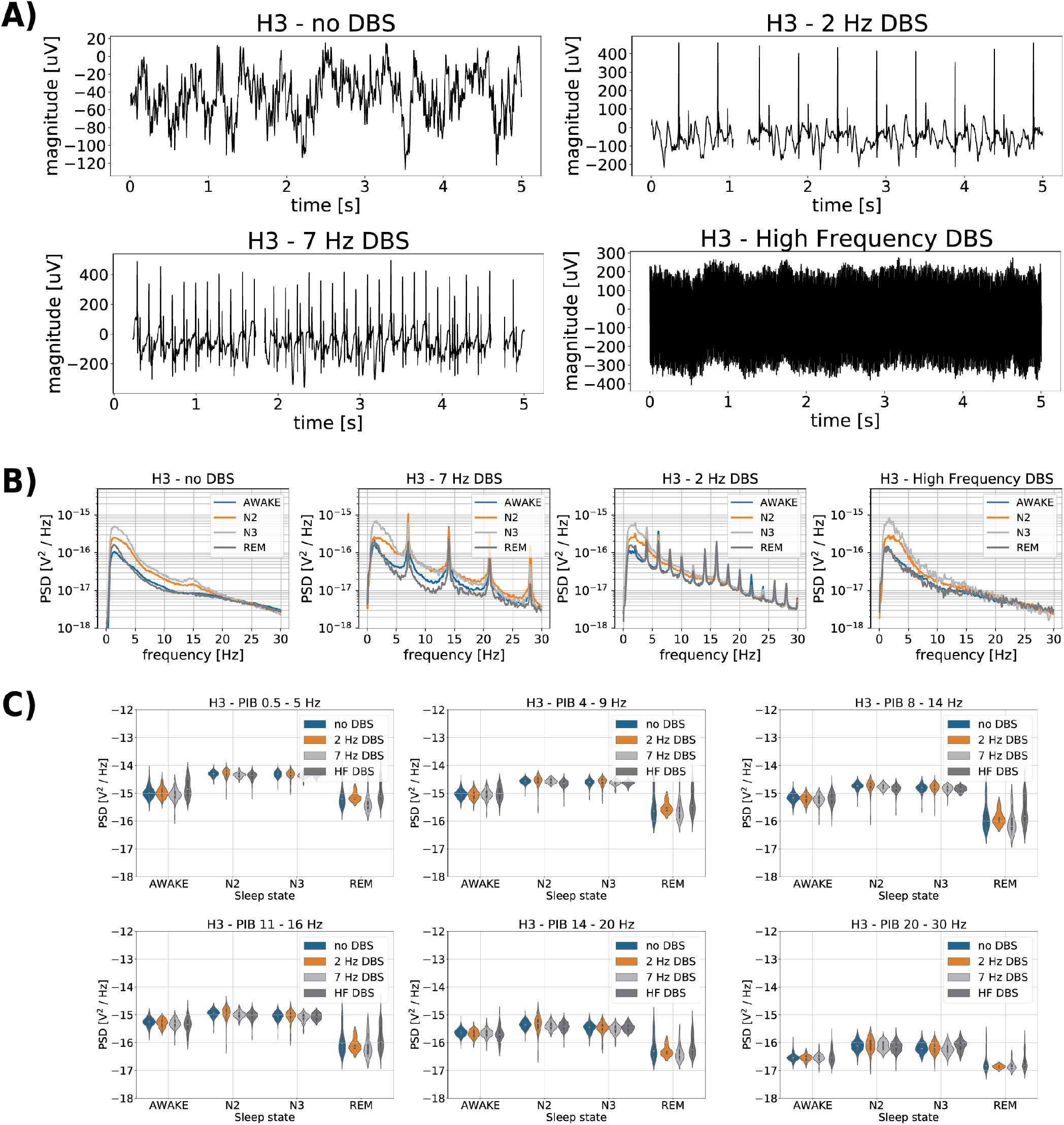
The effect of ANT DBS on iEEG signals. **A)** The iEEG recorded from right hippocampus (HPC) (patient H3) for different ANT DBS frequencies (No DBS, 2, 7 Hz and HF DBS). **B)** Power spectral density (PSD) for different ANT DBS frequencies (No DBS, 2 Hz, 7 Hz and HF DBS): average of each sleep phase spectrum across three nights using 30-second window, estimated by Welch’s method (Awake, N2, N3, REM); **C)** Power in band features (0.5 - 5 Hz; 4 – 9 Hz; 8 – 14 Hz; 11 – 16 Hz, 14 – 20 Hz, 20 – 30 Hz) extracted from raw HPC iEEG signals from each sleep states (Awake, N2, N3, REM), over three nights using 30-second window. These data from subject H3 are representative of all patients. High frequency (HF>100 Hz). Rapid eye-movement (REM) sleep, non-REM (N2, N3) sleep, Anterior nucleus of thalamus (ANT), intracranial EEG (iEEG), Hippocampus (HPC), Deep brain stimulation (DBS), Power spectral density (PSD)

**Supplementary Figure 7.**
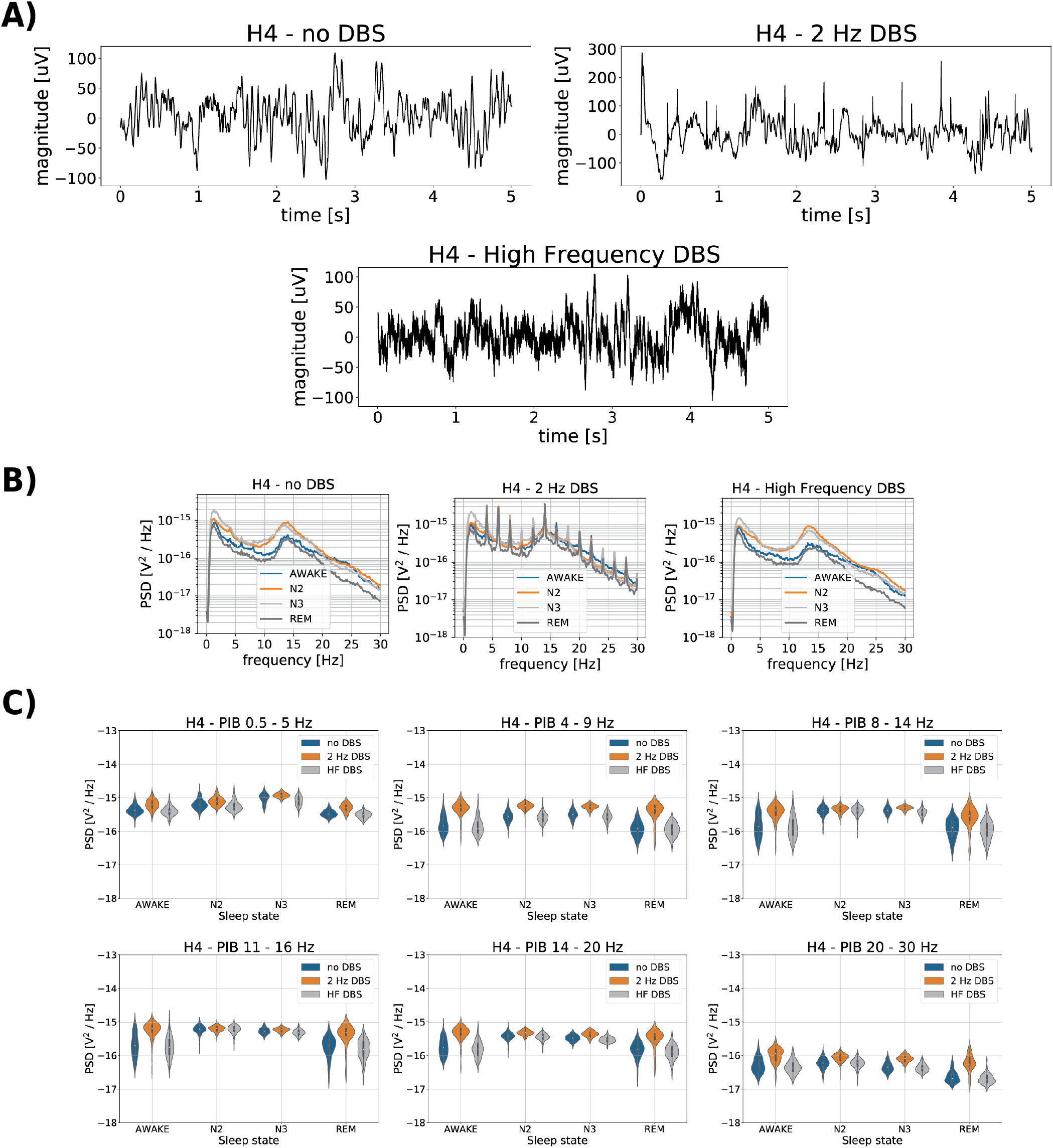
The effect of ANT DBS on iEEG signals. **A)** The iEEG recorded from right hippocampus (HPC) (patient H4) for different ANT DBS frequencies (No DBS, 2 Hz and HF DBS). **B)** Power spectral density (PSD) for different ANT DBS frequencies (No DBS, 2 Hz and HF DBS): average of each sleep phase spectrum across three nights using 30-second window, estimated by Welch’s method (Awake, N2, N3, REM); **C)** Power in band features (0.5 - 5 Hz; 4 – 9 Hz; 8 – 14 Hz; 11 – 16 Hz, 14 – 20 Hz, 20 – 30 Hz) extracted from raw HPC iEEG signals from each sleep states (Awake, N2, N3, REM), over three nights using 30-second window. These data from subject H3 are representative of all patients. High frequency (HF>100 Hz). Rapid eye-movement (REM) sleep, non-REM (N2, N3) sleep, Anterior nucleus of thalamus (ANT), intracranial EEG (iEEG), Hippocampus (HPC), Deep brain stimulation (DBS), Power spectral density (PSD)

**Supplementary Table S3.** *Results of automated behavioral sleep state classification (F1-score) into Awake, REM, N2, N3, sleep categories for all subjects under various settings of electrical deep brain stimulation (DBS) in anterior nucleus of thalamus (ANT). The classification score is reported for the general non-REM category comprising N2 & N3 as well. The feasibility of automated sleep classification using a single channel iEEG data recorded from HPC was performed using no DBS data. We used available iEEG data acquired without DBS during the first night to train the classifier and data acquired during the second and third night for pseudo-prospective (PP) testing. The nights without DBS, however, were not consistent across patients. If data without DBS were not available for the first night, the data without DBS acquired during the second night were used for training and the third night data were utilized for testing. For the 2, 7 Hz & HF DBS, we utilized classifiers trained using no DBS data and transferred the model to the stimulation (MT) data. The experiment was replicated twice, with and without band power cancelling, as introduced in “Cancelling of Band Bower at Stimulation Frequencies”. Moreover, 20 % cross-validation testing (CV) proportionally sampled across all classification categories was performed for the high frequency DBS setups. Values marked by * were achieved using less than 10 samples and therefore are not considered valid*. High frequency (HF>100 Hz). Rapid eye-movement (REM) sleep, non-REM (N2, N3) sleep, Anterior nucleus of thalamus (ANT), intracranial EEG (iEEG), Hippocampus (HPC), Deep brain stimulation (DBS)

| EBS Setup | Validation Method | Band Power Cancelling | Subject | Awake | REM | N2 | N3 | non-REM | All | All non-REM |
| --- | --- | --- | --- | --- | --- | --- | --- | --- | --- | --- |
| No DBS | PP | No | H1 | .946 | .703 | .519 | .723 | .923 | .800 | .910 |
|  |  |  | H2 | .848 | .660 | .740 | .338 | .944 | .691 | .893 |
|  |  |  | H3 | .759 | .497 | .547 | .563 | .803 | .669 | .753 |
|  |  |  | H4 | .798 | .763 | .621 | .763 | .924 | .719 | .890 |
|  |  |  | Average | **.838** | **.656** | **.607** | **.597** | **.899** | **.720** | **.862** |
|  |  | Yes 2 Hz | H1 | .941 | .736 | .415 | .700 | .923 | .775 | .910 |
|  |  |  | H2 | .863 | .712 | .720 | .196 | .944 | .691 | .896 |
|  |  |  | H3 | .677 | .323 | .074 | .488 | .748 | .654 | .736 |
|  |  |  | H4 | .246 | .000 | .265 | .204 | .803 | .576 | .790 |
|  |  |  | Average | **.682** | **.443** | **.369** | **.397** | **.855** | **.674** | **.833** |
|  |  | Yes 7 Hz | H1 | .934 | .613 | .333 | .709 | .935 | .779 | .904 |
|  |  |  | H2 | .844 | .712 | .774 | .360 | .952 | .733 | .909 |
|  |  |  | H3 | .732 | .503 | .597 | .613 | .812 | .666 | .744 |
|  |  |  | H4 | .829 | .776 | .533 | .605 | .899 | .704 | .879 |
|  |  |  | Average | **.835** | **.651** | **.484** | **.572** | **.900** | **.721** | **.859** |
| 2 Hz | MT | No | H1 | .991 | .753 | .455 | .750 | .897 | .765 | .892 |
|  |  |  | H2 | .960 | .714 | .552 | .440 | .960 | .755 | .940 |
|  |  |  | H3 | .383 | .544 | .804 | .714 | .920 | .676 | .763 |
|  |  |  | H4 | .696 | .491 | .169 | .522 | .788 | .480 | .781 |
|  |  |  | Average | **.758** | **.626** | **.495** | **.607** | **.891** | **.669** | **.844** |
|  |  | Yes 2 Hz | H1 | .991 | .848 | .364 | .721 | .932 | .789 | .931 |
|  |  |  | H2 | .955 | .727 | .437 | .442 | .932 | .743 | .936 |
|  |  |  | H3 | .783 | .426 | .442 | .712 | .883 | .683 | .817 |
|  |  |  | H4 | .286 | .111 | .127 | .072 | .763 | .525 | .768 |
|  |  |  | Average | **.754** | **.528** | **.343** | **.487** | **.878** | **.685** | **.863** |
| 7 Hz | MT | No | H1 | .977 | .100* | .250 | .635 | .929 | .756 | .926 |
|  |  |  | H2 | .964 | .643 | .446 | .295 | .973 | .787 | .957 |
|  |  |  | H3 | .818 | .700 | .774 | .750 | .879 | .807 | .845 |
|  |  |  | H4 | - | - | - | - | - | - | - |
|  |  |  | Average | **.920** | **.672** | **.490** | **.560** | **.927** | **.783** | **.909** |
|  |  | Yes 7 Hz | H1 | .974 | .353* | .444 | .690 | .949 | .815 | .941 |
|  |  |  | H2 | .976 | .914 | .396 | .491 | .978 | .829 | .973 |
|  |  |  | H3 | .870 | .903 | .886 | .810 | .935 | .884 | .918 |
|  |  |  | H4 | - | - | - | - | - | - | - |
|  |  |  | Average | **.940** | **.909** | **.575** | **.664** | **.954** | **.843** | **.944** |
| **High**<br>**Frequency**<br>**> 100 Hz** | MT | No | **H1** | .948 | .767 | .381 | .689 | .950 | .768 | .920 |
|  |  |  | **H2** | .506 | .667 | .522 | .250 | .896 | .658 | .814 |
|  |  |  | **H3** | .458 | .353* | .478 | .571* | .761 | .574 | .652 |
|  |  |  | **H4** | .774 | .800 | .841 | .673 | .971 | .857 | .939 |
|  |  |  | **Average** | **.672** | **.745** | **.556** | **.537** | **.895** | **.714** | **.831** |
|  | CV | No | **H1** | .970 | .894 | .564 | .771 | .987 | .850 | .975 |
|  |  |  | **H2** | .703 | .772 | .613 | .176 | .944 | .772 | .902 |
|  |  |  | **H3** | .899 | .716* | .693 | .000* | .857 | .826 | .899 |
|  |  |  | **H4** | .767 | .702 | .926 | .725 | .975 | .895 | .926 |
|  |  |  | **Average** | **.835** | **.789** | **.699** | **.418** | **.941** | **.836** | **.926** |

## Notes

### Clinical Trial

https://clinicaltrials.gov/ct2/show/NCT03946618

### Author Declarations

This human subjects research study was carried out under an FDA IDE: G180224 and Mayo Clinic IRB: 18-005483 "Human Safety and Feasibility Study of Neurophysiologically Based Brain State Tracking and Modulation in Focal Epilepsy".

